# Reducing Domain Shift For Mitosis Detection Using Preprocessing Homogenizers

**DOI:** 10.1101/2021.09.02.21263039

**Authors:** Sahar Almahfouz Nasser, Nikhil Cherian Kurian, Amit Sethi

**Affiliations:** MeDAL Lab, Electrical Engineering, Indian Institute of Technology Bombay, India

**Keywords:** Domain Generalization, Mitotic Figures, Histopathology, Homogenizers

## Abstract

The detection of mitotic figures in histological tumor images plays a vital role in the decision-making of the appropriate therapy. However, tissue preparation and image acquisition methods degrade the performances of the deep learning-based approaches for mitotic figures detection. MItosis DOmain Generalization challenge addresses the domain-shift problem of this detection task. This work presents our approach based on preprocessing homogenizers to tackling this problem.

## Introduction

Machine learning algorithms often underperform when they are validated on an external data that differs significantly from the distribution of their training data. This problem is even more pronounced in medical imaging modalities due to several intrinsic sources that can contribute to this variability. The MIDOG challenge presents the problem of domain shift in data from the perspective of mitosis detection on large cohorts of histopathology dataset collected from several scanners. Mitosis detection by itself is a very challenging problem owing to the large variability in the morphology of mitosis nuclei, along with the presence of several imposter nuclei. This difficulty is cemented by the large scale inter-observer variabilities. In an endeavour to reduce the domain discrepancy, we present a preprocessing pipeline that acts as an unsupervised domain generalizer that averages the appearance between the different scanners with an additional capability to nullify domain specific signals. This deep learning pipeline leverages the property of auto-encoders as a cross-data homogenizer, essentially reducing the appearance between the different domains [1]

## Material and Methods

Our algorithm was trained on MItosis DOmain Generalization (MIDOG) dataset only.The algorithm consists of a homogenizer followed by RetinaNet [2].

### Dataset

The MIDOG challenge presented samples obtained from four slide scanners systems namely the Hamamatsu XR NanoZoomer 2.0, the Hamamatsu S360, the Aperio ScanScope CS2 and the Leica GT450. Around 50 scans were provided from each of these scanner. The entire training data hence consisted of 200 Whole Slide Images (WSIs) from human breast cancer tissue samples stained with routine Hematoxylin & Eosin (H&E) dye. Supervised training annotations were provided for three scanners except the LeicaGT450. The supervision consisted of mitotic figures and hard negatives that resembles mitotic nuclei. Annotations were collected from multiple experts who were blinded to one another. The preliminary test set contains five WSIs correspond to four unrevealed scanners of which only two were also part of the training set. Evaluating the algorithms was accomplished based on this preliminary test before publishing preliminary results on a leaderboard. The final test contains 20 WSIs belong to the same four scanners used for the preliminary test set.

### Methodology

As mentioned, the object detection using RetinaNet remains unaltered, whereas our main contribution lies in inventing a domain generalising preprocessing step. The preprocessing pipeline consists of a multihead encoder network *G*_*f*_. The encoder coupled with a decoder component *G*_*r*_ completes the autoencoder section of the pipeline that tries to reconstruct the input images, ***x*** ∈ *X* with an L2 loss. This optimization in mean square loss sense results in reconstructing images that hold an average appearance to all the variability in the training images. Utilizing this idea, we use the whole dataset, with the appropriate validation splits, provided as a part of MIDOG challenge in order to make the autoencoder learn all the latent domains present in data. This is feasible as this part of the training does not require an associated supervised label.

The encoder network also has a training adversarial head *G*_*y*_ which basically acts as a domain discriminator. The reason for incorporating this module is to further erase domain specific signals explicitly with the help of an adversarial component. Here the training process makes use of explicit domain labels in the form of the scanner technology labels, ***y*** ∈ *Y* = {*HamamatsuRx, HamamatsuS*360, *Aperio, LeicaGT*450}. We have summarized the overall architecture in Fig Fig. 1. Mathematically, if we denote the output of the encoder is a D-dimensional feature vector ***f***, For every input ***x***, the outputs of the model are the reconstructed image ***r*** and the domain label ***y***.

**Fig. 1.**
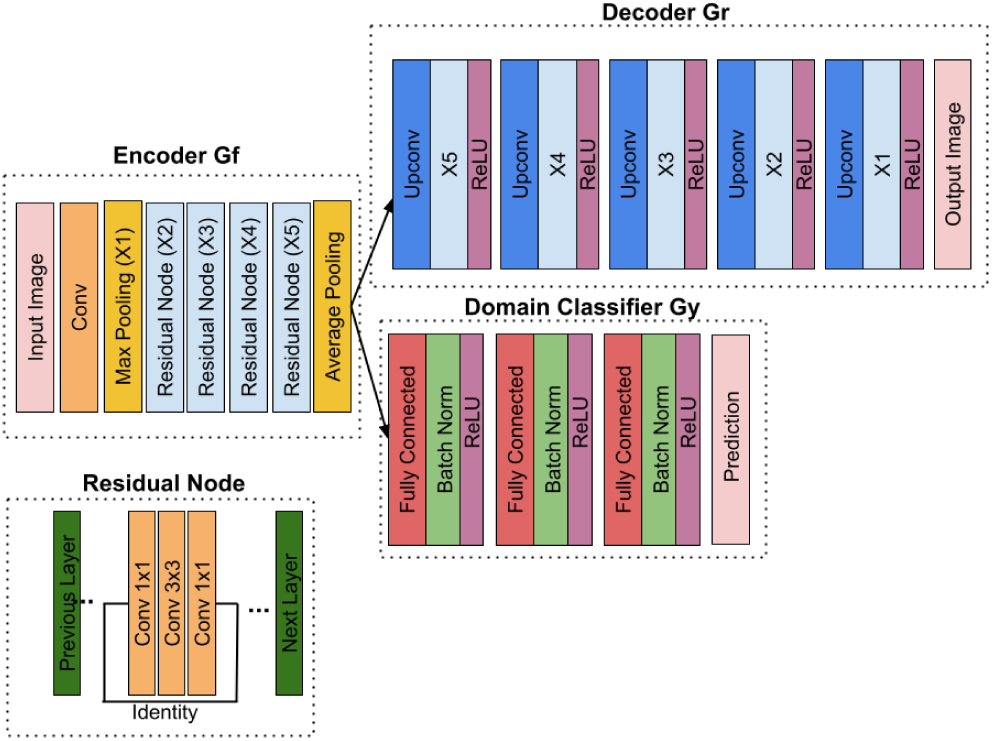
The architecture of the homogenizer

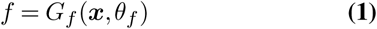

Finally, during the learning stage, we aim to maximize the domain label prediction loss and minimize the reconstruction loss simultaneously to obtain domain-invariant features.

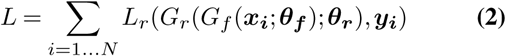

### Network Training

From the data of each of the four scanners we selected 40 WSIs for training the homogenizer and 10 WSIs for validation WSIs. We used a patch size of 256 × 256 pixels and a batch size of 8. Furthermore, we performed data augmentation with ColorJitter, affine transformations and random lightning and contrast change. We trained the network with a cyclical maximal learning rate of 10^−4^ for 60 epochs until convergence. The loss of the homogenizer is a weighted combination of the perceptual loss and the classification loss. And the model optimized by minimizing the reconstruction loss and maximizing the domain classification loss. For object detection, we followed the same strategy of splitting the data of each of the three annotated scanners into 40 WSIs for training and 10 WSIs for validation. We used the focal loss as the classification Loss [3] and L1 loss for regression. We trained the network with a learning rate of 10^−4^ for 150 epochs until convergence.

## Discussion and Conclusion

In this paper, we have described our method for the MIDOG challenge [4]. Our code will be made publicly available in our GitHub repository after the final submission deadline.

## Data Availability

https://imi.thi.de/midog/

